# A COVID-19 Nursing Home Transmission Study: sequence and metadata from weekly testing in an extensive nursing home outbreak

**DOI:** 10.1101/2020.09.15.20195396

**Authors:** Judith H. van den Besselaar, Reina S. Sikkema, Fleur M.H.P.A Koene, Laura W. van Buul, Bas B. Oude Munnink, Ine Frénay, René te Witt, Marion P.G. Koopmans, Cees M.P.M. Hertogh, Bianca M. Buurman

**Affiliations:** Department of Internal Medicine, section of Geriatric Medicine, Amsterdam Public Health Research Institute, Amsterdam University Medical Center, Amsterdam, the Netherlands; Department of Viroscience, Erasmus Medical Center, Rotterdam, the Netherlands; Department of Medical Microbiology, Amsterdam University Medical Center, Amsterdam, the Netherlands; Department of Infectious Diseases, Public Health Laboratory, Public Health Service of Amsterdam, Amsterdam, the Netherlands; Department of Medicine for Older People, Amsterdam Public Health Research Institute, Amsterdam University Medical Center, Amsterdam, the Netherlands; Regional Laboratory for Medical microbiology (RLM) Dordrecht- Gorinchem, Dordrecht, the Netherlands; Eurofins | NMDL-LCPL, Rijswijk, The Netherlands

## Abstract

**Background:** This study aimed to assess the contribution of asymptomatic and presymptomatic residents and staff in SARS-CoV-2 transmission during a large outbreak in a Dutch nursing home.

**Methods:** Observational study in a 185-bed nursing home with two consecutive testing strategies: testing of symptomatic cases only, and weekly facility-wide testing of staff and residents regardless of symptoms. Nasopharyngeal and oropharyngeal testing with RT-PCR for SARs-CoV-2 was conducted with a standardized symptom assessment. Positive samples with a cycle threshold (CT) value below 32 were selected for sequencing.

**Results:** 185 residents and 244 staff participated. Sequencing identified one cluster. In the symptom-based test strategy period 3/39 residents were presymptomatic versus 38/74 residents in the period of weekly facility-wide testing (p-value<0.001). In total, 51/59 (91.1%) of SARS-CoV-2 positive staff was symptomatic, with no difference between both testing strategies (p-value 0.763). Loss of smell and taste, sore throat, headache or myalga was hardly reported in residents compared to staff (p-value <0.001). Median Ct-value of presymptomatic residents was 21.3, which did not differ from symptomatic (20.8) or asymptomatic (20.5) residents (p-value 0.624).

**Conclusions:** The frequency of a/presymptomatic residents compared to staff suggests that a/presymptomatic residents could be unrecognized symptomatic cases. However, symptomatic and presymptomatic/unrecognized symptomatic residents both have the same potential for viral shedding. The high prevalence symptomatic staff found in facility-wide testing suggests that staff has difficulty attributing their symptoms to possible SARS-CoV-2 infection. Weekly testing was an effective strategy for early identification of SARS-Cov-2 cases, resulting in fast isolation and mitigation of this outbreak.

## INTRODUCTION

Worldwide, nursing homes (NHs) are facing outbreaks of severe acute respiratory syndrome coronavirus 2 (SARS-CoV-2) with high case fatality rates.^1,2^ In the Netherlands, at the end of July there had been outbreaks of COVID-19 in 1052 of the 2500 NHs, with in total 8433 confirmed cases and 2850 deaths.^3^ The current CDC-guideline and ECDC guideline recommend expanded viral testing of asymptomatic residents in NHs if a single new case of a SARS-CoV-2 infection is detected, based on data of previous NH outbreaks which suggest an important role for presymptomatic spread of SARS-COV-2 among residents^4-10^. However, it remains unknown to which extent asymptomatic and presymptomatic cases contribute to the spread of SARS-CoV-2. Also, specifically in the NH setting, it remains unclear to what extent asymptomatic cases are truly without symptoms. Sole reliance on symptoms for testing in NHs could be insufficient because self-reporting of complaints is often compromised in residents due to limited ability to communicate (e.g. in residents with dementia).^11^ The Dutch guideline for COVID-19 in NHs states that only residents with possible symptoms of SARS-CoV-2 should be tested^12^ and no policy for testing of asymptomatic residents or staff is facilitated in the Netherlands.

Multiple reports have been published about the prevalence of asymptomatic and presymptomatic residents and staff in NHs after the implementation of a facility-wide testing strategy during an outbreak.^5,7,10,13,14^ The prevalence of asymptomatic staff and residents differed from single cases to up to half of the infected cases. Low cycle threshold (Ct) values were found in asymptomatic and presymptomatic cases, suggesting potential of viral shedding.^7,10^ A large registry of 857 Dutch residents with confirmed SARS-CoV-2 showed that 93% of cases expressed any of the symptoms cough, shortness of breath or fever. A large range of other symptoms were also reported: 21% of cases expressed fatigue and in 10-15% of cases symptoms such as diminished intake, gastro-intestinal symptoms, malaise and rhinorrhea occurred.^15^ However, the presentation of SARS-CoV-2 can be difficult to recognize in NH residents, which can cause delay in testing, isolation and treatment.^15,16^ In addition, during a community-wide outbreak it can be difficult to distinguish residential outbreaks from multiple introductions without sequencing of viruses from cases.^17^

Viral spread by presymptomatic or unrecognized symptomatic cases has important implications for Personal Protective Equipment (PPE) use, facility-wide testing and isolation measures in NHs for the prevention of outbreaks. The aim of this study is to analyze the contribution of presymptomatic spread of SARS-CoV-2 in all staff and residents of a NH in the Netherlands by serial weekly point prevalence surveys, PCR and sequencing.

## METHODS

### Setting and study population

The study took place in a 185-bed NH in the province South Holland which provides long-term care and is specialized in dementia care. All residents and staff working during the outbreak were invited to participate in the study. Data was collected retrospectively before May 18^th^ and prospectively from May 18th onwards. NH details are presented in ***Supplementary Material 1***.

### SARS-CoV-2 testing and analysis

Two phases in the NH test strategy can be distinguished: First, until May 11^th^, a symptom-based testing strategy was followed, according to national guidelines: cases were tested when they experienced any symptoms. The only exception of this strategy was at May 6th: at the ward where the outbreak started, all negative residents were tested regardless of symptoms. Second, from May 12^th^ the NH implemented a policy of facility-wide weekly testing in addition to the symptom-based testing strategy, implying SARS-CoV-2 testing of all residents without a previous positive test and regardless of the presence of any symptoms. Staff was tested regardless of symptoms in the week of May 18^th^ and June 1^st^.

Samples were transported to collaborating laboratories at the end of each test day, where they were tested for SARS-CoV-2 polymerase chain reaction (PCR) targets. Three different laboratories collaborated because of the large number of tests which were conducted: As a result, different PCR platforms were used, however the used targets were similar (RdRp-gene, E-gene, N-gene) see ***Supplementary material 2***.

### Sequencing, Phylogenetic analysis and cluster definition

PCR-positive samples with Ct-value below 32 were selected for sequencing using a SARS-CoV-2 specific amplicon based Nanopore sequencing approach, as previously described.^18^ The consensus genome was generated only including positions with a coverage >30 as described previously.^19^

Sequences with >10% “Ns” were excluded and compared to a reference database developed for the national COVID-19 response effort ***(Supplementary Material 3)***.^20^ The alignment was manually checked for discrepancies after which IQ-TREE^21^ was used to perform a maximum likelihood phylogenetic analysis under the GTR+F+I⍰+G4 model as best predicted model using the ultrafast bootstrap option with 1,000 replicates. The phylogenetic trees were visualized in Figtree.^22^ For clarity reasons all bootstrap values below 80 were removed.

### Data collection

A standardized symptom-assessment form was completed by the research team for each assenting resident, using electronic health record review. Staff was invited to complete a first questionnaire electronically (via email) in the week of May 18^th^. ***(Supplementary Material 4 and 5)***

The standardized symptom-assessment included fever (defined as temperature greater than 38.0°C (100.4°F)), cough, shortness of breath, chills, malaise, fatigue, rhinorrhea, nasal congestion, sore throat, myalgia, headache, nausea or diarrhea, diminished intake, and loss of smell or taste. For residents, increased confusion and decreased oxygen saturation was registered additionally. A participant was classified symptomatic if he had at least one new or worsened symptom in the 14 days prior to a positive test result (T0). A participant was classified asymptomatic if no new or worsened symptoms were present and no symptoms would develop in the fourteen days following the positive test. Participants were classified pre-symptomatic if they had no symptoms at moment of testing, but developed symptoms in the two weeks following a positive test (T1, T2).

### Analyses

Baseline characteristics were analyzed descriptively. Participants were characterized with means, medians, range and standard deviations (SD) for continuous variables and counts with percentages for categorical data. Differences between groups were assessed with student’s T-test, Mann-Whitney U and Kruskal-Wallis test for continuous variables and Chi-Square test for categorical data. Differences were considered statistically significant at P <0.05 (2-tailed). All analyses were done using SPSS, version 26 (IBM, Armonk, NY) and Excel.

### Ethics

Written information about the study was sent out to residents and their legal representatives at May 18^th^, with the possibility to opt-out. Health care professionals were asked informed consent for participating in the study prior to digital questionnaire completion. The Medical Ethics Committee of the VU University Medical Centre in Amsterdam reviewed the study protocol and confirmed that the study does not fall under the scope of the Medical Research Involving Human Subjects Act.

## RESULTS

At April 29^th^, when the first resident tested positive for SARS-CoV-2, 185 residents lived and 384 staff worked in the NH. Four legal representatives of residents and 34 staff members declined participation. Baseline characteristics are described in ***Table 1***. Residents who tested positive for SARS-CoV-2 were older and more likely to have cognitive impairment. Staff positive for SARS-CoV-2 consisted mostly of (registered) health care assistants and health-care aids. ***Supplementary Material 6*** shows the STROBE diagram of participating residents and staff.

**Table 1:** Baseline characteristics residents and staff.

|  | SARS-CoV-2 Test Results |  | p-value, (95% CI interval) |
| --- | --- | --- | --- |
| **Residents** | **Positive**<br>**N=113** | **Negative**<br>**N=68** |  |
| Age (median/ range) | 85.0 (44-99) | 81.5 (48-100) | 0.001 (-9.009 ; -2.237) |
| Female n. (%) | 82 (72.6) | 50 (73.5) | 0.888 |
| <u>Coexisting conditions</u> |  |  |  |
| Pulmonary disease n(%) | 12 (10.6) | 3 (4.4) | 0.142 |
| Cardiovascular disease n(%) | 40 (35.4) | 15 (22.1) | 0.059 |
| Cerebrovascular disease n(%) | 23 (20.4) | 9 (13.2) | 0.224 |
| Diabetes n(%) | 18 (15.9) | 18 (26.5) | 0.085 |
| Cognitive impairment n(%) | 104 (92.0) | 53 (77.9) | 0.007 |
| Reduced kidney function n(%) | 7 (6.2) | 2 (2.9) | 0.329 |
| Obesity n(%) | 5 (4.4) | 8 (11.8) | 0.064 |
| **Staff\*** | **Positive**<br>**N=56** | **Negative**<br>**N=188** |  |
| Age (median/ range) | 43.0 (18-74) | 46.5 (18-74) | 0.853 (-5.764 – 3.942) |
| Female n (%) | 47 (83.9) | 175 (93.1) | 0.036 |
| <u>Profession, n (%)</u> |  |  | 0.027 |
| Health care assistants and aids | 39 (69.6) | 88 (46.8) |  |
| Nurse | 3 (5.4) | 11 (5.9) |  |
| Physical therapist | 0 | 7 (3.7) |  |
| Physician | 0 | 6 (3.2) |  |
| Other** | 14 (24.6) | 76 (40.4) |  |
| <u>Reporting contact with Covid-19 suspected or confirmed residents, n(%)</u> |  |  | 0.296 |
| Yes | 43 (76.8) | 159 (84.6) |  |
| No | 5 (8.8) | 8 (4.3) |  |
| Unknown | 8 (14.0) | 21 (11.2) |  |

\* 34 Staff members declined participation, 106 staff did not complete the questionnaire
\*\* Staff working in kitchen, logistics, occupational therapists, psychologists, management

### Introduction of the virus and outbreak

The first positive resident (29 April) had been admitted from 17-23 April at the geriatric department of the local hospital with a urosepsis. She had a negative PCR for SARS-CoV-2 and her Chest X-Ray was classified as CORADS-1, suggesting a very low probability of COVID-19. April 29^th^ she developed a fever and was readmitted to the hospital, where retrospectively an outbreak had occurred, and tested positive for SARS-CoV-2. Previously, three NH staff members tested positive for SARS-CoV-2 in April, but none of them worked in the period they were contagious.

### Sequencing

In total, 53 sequences of clients and NH employees were available. In addition, 7 sequences of the hospital outbreak were generated. All sequences cluster together, also sequences detected at the geriatric department of the hospital outbreak were near identical. Two subclusters appear to be present in sequences of clients and employees, without differences when considering wards where residents lived and employees worked. ***(Figure 2)***

**Figure 1:**
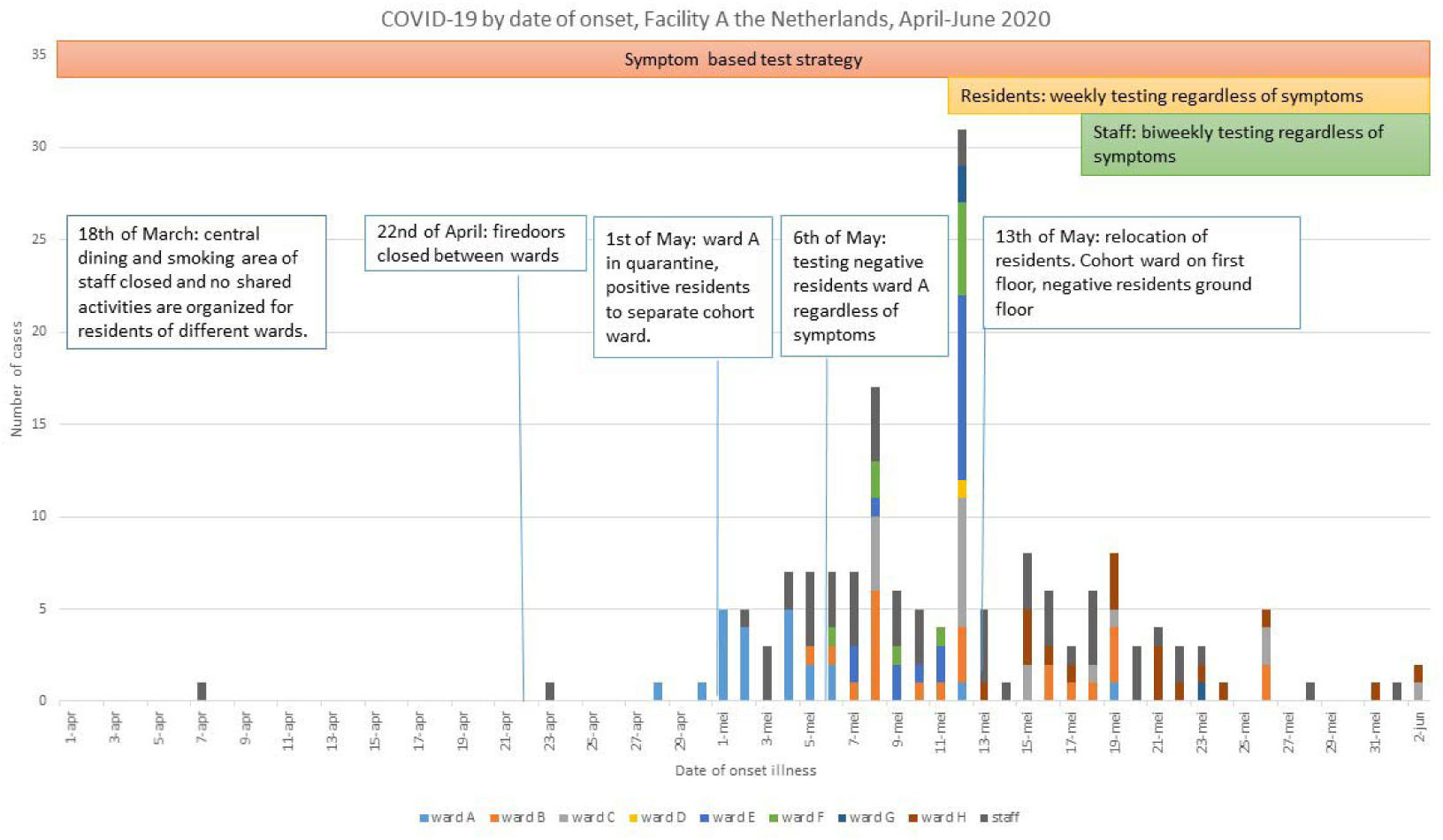
COVID-19 by date of onset and nursing home policy. Figure 1 shows the date of onset of COVID-19 for participating residents of the different wards and participating staff from the 15^th^ of April until the 2 ^nd^ of June. Key changes in NH policy for infection prevention and testing are indicated. On May 13^th^, facility management decided to move all positive tested residents to the first floor of the building, while residents who tested negative were moved to the ground floor of the building. PPE used on the first floor included isolation gown, gloves over the wrists, goggles and a surgical mask; on the ground floor surgical masks and gloves were used.

**Figure 2:**
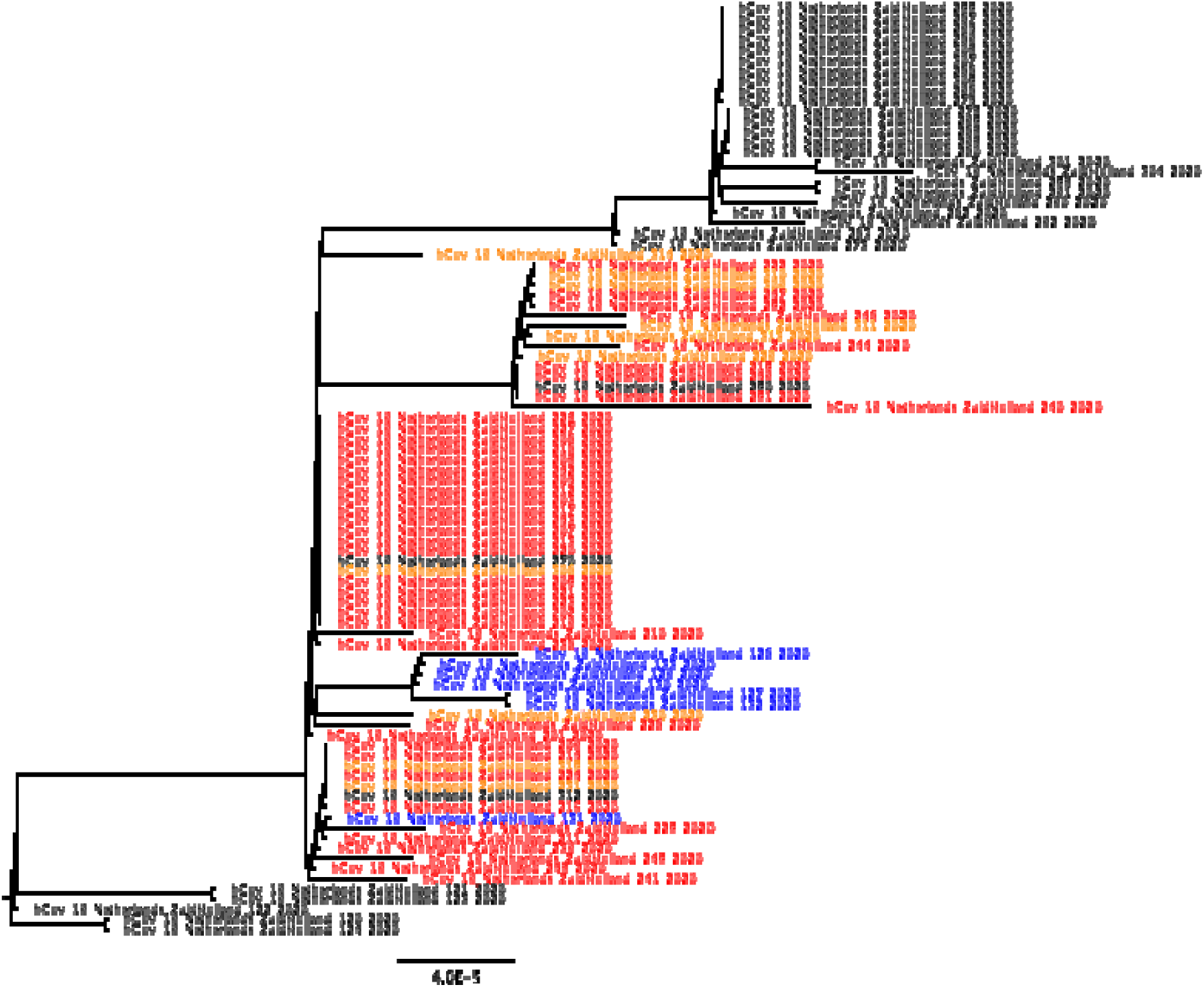
Zoom-in of Dutch phylogenetic tree, with sequences of nursing home A in red (clients) and orange (employees). Sequences in blue originate from the related hospital outbreak. Sequences in black originate from a Dutch national reference database.

**Figure 4:**
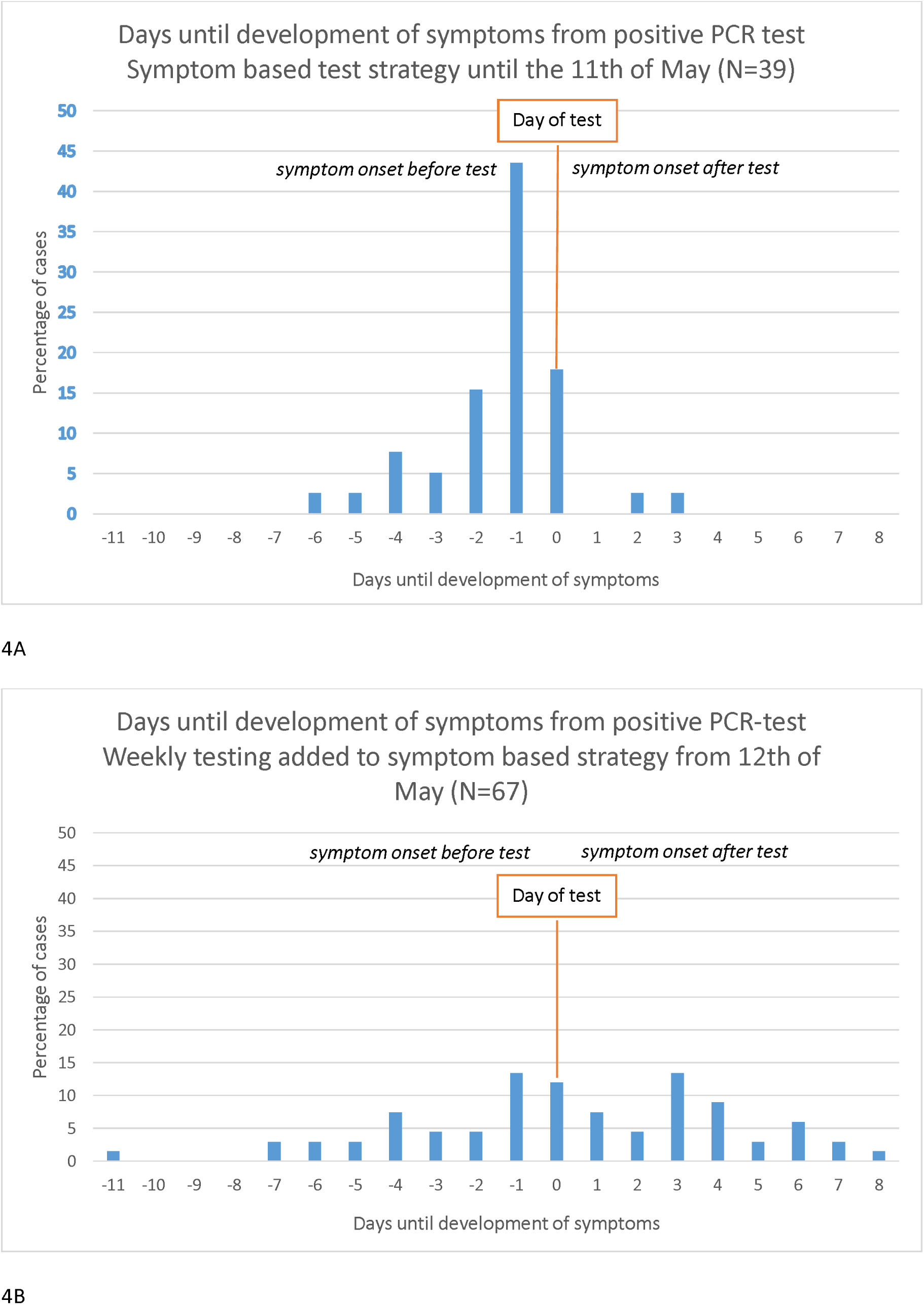
frequency plot of days until development of symptoms from positive PCR-test of residents. Negative values represent symptomatic residents, while positive values represent presymptomatic residents. The value 0 means that residents developed symptoms at the day of PCR-test: whether the symptoms developed before or after testing determines if they were presymptomatic or symptomatic. A) symptomatic testing strategy until the 11^th^ of May. B) Addition of facility-wide weekly testing strategy regardless of symptoms from the 12^th^ of May.

### Symptomatic, presymptomatic and asymptomatic cases during symptomatic and weekly testing strategy

Results of the standardized symptom-assessment are presented in ***Table 2***. Except for the symptoms fever and nausea, residents and staff showed different prevalence for all symptoms.

**Table 2:**
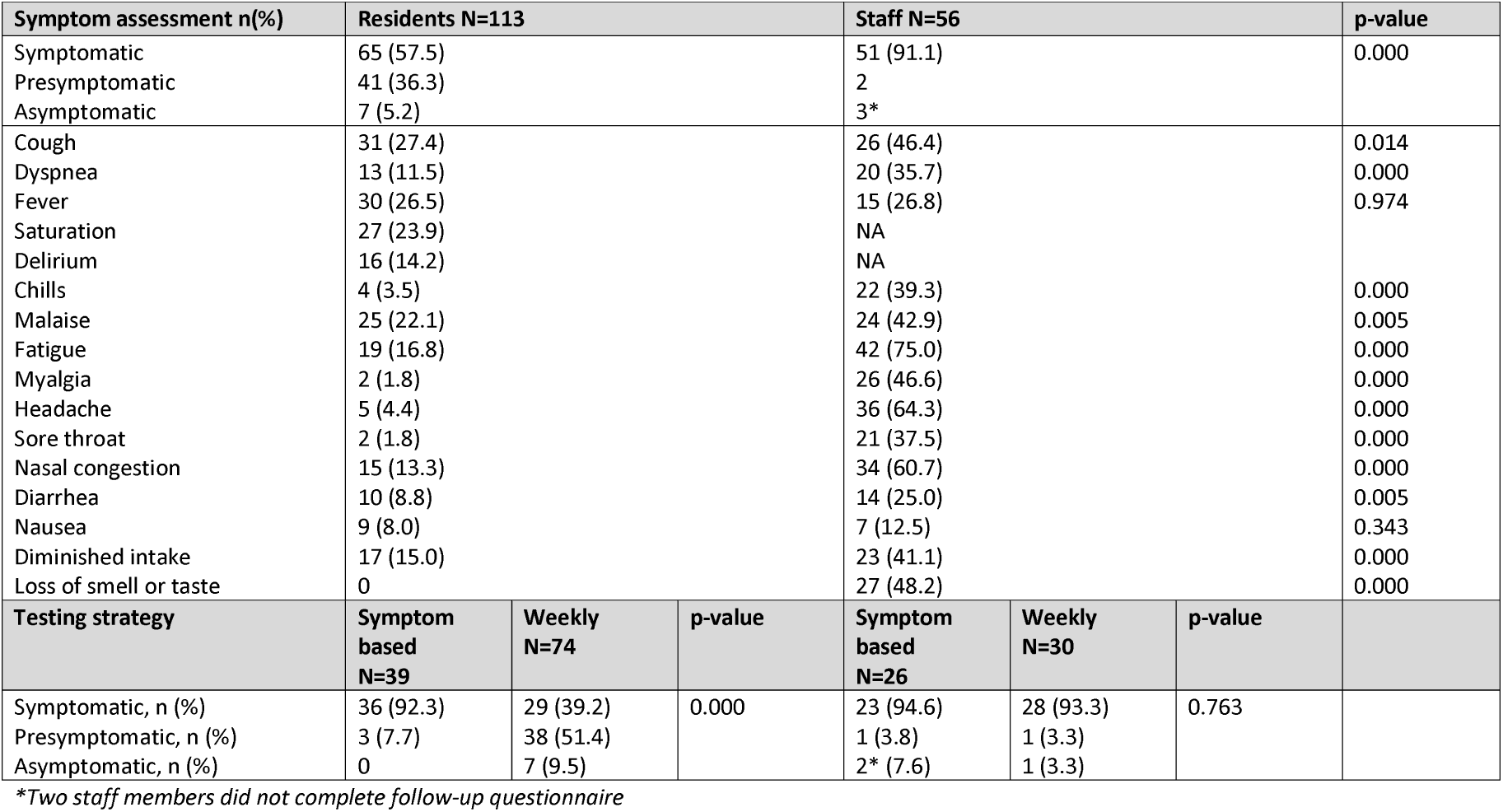
Symptom assessment of residents and staff with a positive SARS-CoV-2 PCR-test.

A significant difference in presymptomatic residents was found between the two testing strategies (p-value <0.001). Before the start of facility-wide weekly testing, 39 residents tested positive: 36 (92.3%) were symptomatic and 3 (7.7%) residents were presymptomatic. The three presymptomatic residents were tested at May 6^th^ when all residents of the ward where the outbreak started were tested regardless of symptoms. In the period of weekly testing, 74 residents tested positive, of which 29 (39.2%) were symptomatic at the time of testing, 38 (51.4%) were presymptomatic and 7(9.5%) were asymptomatic.

A total of 56 staff tested positive and completed the questionnaire: 51 (91.1%) were symptomatic at the moment of testing, 2 (3.9%) were pre-symptomatic and 3 (5.9%) staff members were asymptomatic. No difference in symptomatic, presymptomatic and asymptomatic staff members was found between symptom based or additional weekly testing strategy (p-value 0.763).

### Symptom onset and presentation with symptomatic and weekly testing strategy

Until May 11^th^, 39 residents tested positive and all developed symptoms. Symptoms developed 6 days before testing until 3 days after testing, with a median of development of symptoms the day before the test (interquartile range 2 days to 1 day before test). (Figure 4A) After the addition of weekly testing regardless of symptoms, 74 residents tested positive of which 67 residents developed symptoms from 11 days before testing until 8 days after testing, with a median of development of symptoms the day of the test (interquartile range 2 days before the test to 3 days after the test) (Figure 4B). The time between onset of symptoms and test date differed significantly between the two testing strategies (p-value 0.000). With both test strategies symptomatic residents had symptoms for multiple days without testing.

### Ct-values

Ct values were available for 97/113 positive residents; the median Ct-value was 21.3 (range 14.5-40). Symptomatic residents (N=59) had a median Ct-value of 20.8 (range 14.5-38.1), presymptomatic resident (N=33) had a median Ct-value of 21.3 (range 16.1 -40) and asymptomatic resident (N=5) had a median Ct-value of 20.5 (range 17.3-39.7). There was no difference in Ct-value between these groups(P=0.624).

Ct-values were available for 38/56 staff members; with a median of 24.6 (range 13.7-38.1). Of one asymptomatic staff Ct-value was 34.6. The two presymptomatic staff members Ct-values were 29.8 and 32.3. Symptomatic staff members (N=35) had a median Ct-value of 23.7 (range 13.7-38.1).

## DISCUSSION

We describe a large SARS-CoV-2-outbreak in a NH which most likely started by an infected resident discharged from a local hospital were SARS-CoV-2 prevailed. The addition of weekly facility-wide testing regardless of symptoms identified 38 (52.7%) presymptomatic residents and 7 (8.1%) asymptomatic residents. These cases were found up to eight days before symptoms occurred. In staff limited presymptomatic and asymptomatic cases were identified. The absence of subjective symptoms (such as loss of smell or taste) in residents compared to staff who are infected by the same SARS-CoV-2 strain suggests the under-reporting of symptoms in residents. As such, it is not possible to make a distinction between a/presymptomatic and unrecognized symptomatic residents in this study. However, a/presymptomatic residents have the same high viral load as symptomatic residents, which suggests the same potential for viral shedding. This implicates only a symptomatic testing policy during an outbreak is insufficient in NHs.

The high prevalence of presymptomatic cases in residents and the limited registration of subjective symptoms in residents such as headache or a sore throat is comparable to other studies.^4,6,7,10^ In our study, especially loss of smell and taste and a sore throat was hardly reported in residents, while these symptoms were frequent in staff. Studies performing facility wide testing regardless of symptoms of staff in NHs with a confirmed COVID-19 case found the same limited number of asymptomatic staff as in our study.^4,5,14^ We are the first study reporting on symptoms from residents and staff in a large outbreak of the same virus strain. The large difference between presymptomatic staff and residents found in this study has three possible explanations: First, a large number of residents in this NH is cognitive impaired, which makes it difficult for them to express their symptoms. Second, staff reporting on residents’ symptoms are not aware of all the symptoms related to COVID-19. During the outbreak symptoms of residents were sometimes documented for multiple days, but they were nevertheless not tested. Third, understaffing because of the outbreak could have led to suboptimal symptom registration: mild or subjective symptoms were missed, because staff had to take care of residents they were not familiar to work with, or because of limited time to register symptoms which interfered with the follow-up of symptoms and early recognition of possible infected patients. Understaffing as a risk for under-recognition of new cases is supported by data of Gorges and Li which shows NHs with at least one case, higher nurse aide^23^ and total nursing hours^23,24^ are associated with a lower probability of experiencing an outbreak and with fewer deaths.

Our study showed no difference between Ct-values of symptomatic, presymptomatic or asymptomatic cases in residents, similar to previous studies.^7,10^ All these studies found the same risk of under-recognition of (mild) symptoms of SARS-CoV-2 and classifying residents as pre- or asymptomatic. However, all these residents should be treated the same: as possibly infectious. These residents should be isolated and personal protective equipment should be used during their care.

The new approach of mass repeated testing, irrespective of symptoms, in skilled nursing facilities has been advocated since May.^25^ After this, studies have been published describing this approach, often resulting in reduced SARS-CoV-2 transmission after the implementation of this testing strategy.^26,27^ However, limited additional cases were found after a weekly testing strategy was implemented in three Dutch NHs after their first cases of SARS-CoV-2.^28^ Possibly, the testing was early in the outbreak and led to rapid isolation, the increased availability of PPE or because cases per capita in the community were very low.^28^ Cases per capita in the community have been identified as the strongest predictor for outbreaks in NHs.^23^

The testing of staff regardless of symptoms is important because previous research showed that health care workers have difficulty in recognizing possible COVID-19 symptoms for themselves: when interviewing staff with a positive SARS-CoV-2 PCR, 17% did not express any symptoms of fever, cough, shortness of breath or a sore throat. Also, 65% reported working while exhibiting symptoms.^29^ This is reflected in our results, as we found that almost none of the staff was asymptomatic at the moment of testing, even after the implementation of a testing strategy regardless of symptoms. Facility-wide testing always has to be based upon applicable infection prevention testing guidelines, starting with residents and staff most at risk of being infected (close contacts) and extending testing from there if necessary.

Implementing sequencing, combined with epidemiological information, is important to understand the extent of intramural transmission versus introductions from the community. In addition, transmission clusters and risk factors for transmission can be identified, that can be used to implement infection prevention measures to prevent further spread. Previous research has shown that whole genome sequencing can generate evidence for transmission routes that would not have been identified with traditional epidemiological investigations.^17,18^

### Limitations

Not all staff members who tested positive participated in the study. In addition, some staff members had to answer the questionnaire retrospectively, which gives the risk of recall bias. Also, the questionnaire did not inquire if staff worked with symptoms.

Further, the difference between symptomatic staff and residents could perhaps be explained by the fact that staff was tested less frequent than residents: residents were tested weekly, but staff was tested biweekly. This may have contributed partly to the higher proportion of symptomatic staff. Last, not all SARS-CoV-2 positive samples were sequenced. However, a lot of time points could be analyzed and they show all the same cluster which makes it unlikely that multiple clusters were circulating in the NH.

### Conclusion

This study suggests that part of the presymptomatic cases in NHs are unrecognized symptomatic cases. Our study supports the guideline of the CDC and ECDC that facility-wide testing of residents and staff needs to be undertaken after the first confirmed SARS-CoV-2 case in the facility. If there is limited viral testing capacity, initial testing of (asymptomatic) close contacts is advised (CDC). This allows identification of possible asymptomatic, presymptomatic cases and unrecognized symptomatic cases and prevent further spread of the virusSequencing should be performed to discriminate ongoing intramural transmission and multiple introductions. ***Box 1*** summarizes the lessons learned during this study.

## Box 1

**lessons learned of SARS-CoV-2 outbreak**

### Lessons learned

#### 1.

**Preparing for an outbreak**

Educate staff about all the possible symptoms of COVID-19:. Take routine temperature and saturation of residents for reference values. Also sufficient staffing and staff dedicated to a few patients is necessary for early recognition of symptoms. Nursing homes should make protocols with a local laboratory so when an outbreak occurs, rapid testing is possible.

#### 2.

**Increasing cases per capita in the population**

When cases per capita in the general population of the area are increasing, staff and visito should wear at least surgical face mask to prevent possible introduction of the virus. Moreover, a resident transferring from another health care facility (hospital or other nursing home) or home should be kept in quarantine for 10 days, even with a negative SARS-CoV-2 PCR.

#### 3.

**During an outbreak**

Recognition of start of possible COVID-19 symptoms is very difficult, especially in residents with dementia. Weekly testing during an outbreak identifies presymptomatic or unrecognized symptomatic residents and makes timely isolation and use of PPE possible. Also staff should be tested weekly, even while using PPE.

## Data Availability

Data is available upon request.

## ACKNOWLEDGEMENTS

We thank all nurses and physician assistants of the participating NH and Public Health Services for their contribution to and/or performance of SARS-CoV-2 testing. We thank the persons from Amsterdam UMC who assisted in questionnaire administration. We thank Ellen Verspuij, Paul van Gennip, Saskia Heugens and Mariëtte van der Spek for their assistance in logistic and administrative aspects of the study. We thank Inge Huijskens of the Regional Laboratory for Medical microbiology (RLM) for her assistance with collecting sequencing data. We thank Anja Schreijer, Menno de Jong, Mariska Petrignani and Joost Wiersinga for their time and discussion for interpreting the data. We thank the laboratories of Regional Laboratory for Medical microbiology (RLM), Eurofins and Sanquin for the performance of SARS-CoV-2 tests.

